# Respondent-Driven Sampling Recruitment Dynamics Among People Who Smoke Drugs in South Africa

**DOI:** 10.64898/2026.09.15.26363109

**Authors:** Hannah Ruth Sullivan, Tara Carney, Frances Ratangee, Nandi Niemand, Danie Theron, C Robert Horsburgh, Laura F White, Robin M Warren, Karen R Jacobson, Samantha Malatesta

## Abstract

**Background:** Respondent-driven sampling (RDS) is a widely used peer-recruitment strategy to study hard-to-reach populations including people who use drugs. South Africa continues to experience increasing illicit substance use alongside high burdens of tuberculosis (TB) and HIV. Little is known about the factors that influence RDS recruitment success in these settings, especially in rural areas.

**Methods:** A total of 750 people who smoke drugs (PWSD) were recruited through RDS in a rural town in the Western Cape, South Africa to estimate TB and HIV prevalence. Participant characteristics were compared by recruitment success, defined as distributing at least one study coupon that resulted in peer enrollment, to identify factors associated with successful recruitment.

Recruitment homophily was assessed to quantify within-group recruitment, and the stabilization of selected participant characteristics across recruitment waves were visualized to assess sample diversity over time.

**Results:** Recruitment success was more common among unemployed participants (p=0.032), those without an incarceration history (p=0.053), current cannabis users (p=0.086), and participants with moderate-to-severe methamphetamine (p=0.019) or methaqualone (p=0.014) use. High within-group recruitment was observed by neighborhood of residence and racial population group. The proportion of males, individuals with incarceration history, and participants with TB or HIV stabilized across recruitment waves, regardless of seed characteristics.

**Conclusions:** Recruitment dynamics in this setting were strongly influenced by social and geographic factors, including neighborhood, population group, incarceration history, and substance use type and severity. Identifying characteristics associated with recruitment success may inform more efficient peer-recruitment strategies, such as targeted coupon distribution. These findings support the use of RDS as an effective approach for engaging PWSD in research.

## Introduction

Traditional recruitment methods present challenges for public health surveillance and research among hard-to-reach populations, including people who use drugs (PWUD). Hard-to-reach populations, often defined by marginalized characteristics or stigmatizing behaviors, have been systematically underrepresented in research due to socioeconomic barriers, historical exclusion from health systems, and concerns about retention in longitudinal studies (1–3). In rural areas, stigma is often confounded by less anonymity or increased social surveillance (4,5). Many marginalized individuals are not captured in standard sampling frameworks, including health clinics or census records, creating additional barriers to participation in research (1,6). Studies that rely on these sampling frames introduce selection bias, limiting participation among hard-to-reach populations and reducing the accuracy of disease burden and risk behavior estimates (1,7). Alternative sampling methods and study designs have been developed to improve the inclusion of hard-to-reach populations in research.

Respondent-driven sampling (RDS) is one such alternative: a peer-referral strategy in which participants recruit members of their social network by distributing study coupons, which are then returned to the study site for enrollment (6). Recruitment begins with a convenience sample of initial participants, termed *seeds*, who then recruit their peers, generating referral chains that extend across successive recruitment waves (6,8). By leveraging existing social networks, RDS enables estimation of key population characteristics and their prevalence in socially marginalized groups that may otherwise not be captured through conventional methods. As a result, RDS has become an increasingly common approach for studying hard-to-reach populations (9).

People who smoke drugs (PWSD) represent one such population, where overlapping structural and health inequities contribute to elevated infectious disease risk. In South Africa, this is compounded by one of the world’s highest dual burdens of tuberculosis (TB) and HIV (10). Rising methamphetamine and methaqualone use over recent years has raised concern about amplified HIV transmission, especially among marginalized Black African communities that are disproportionately affected by both HIV and recent illicit drug use surges (11). PWSD also present with a higher bacterial burden at TB diagnosis compared to people who do not smoke drugs, with drug use being associated with a fourfold increase in non-adherence to TB treatment (4,12,13). Because drug use and disease risk are patterned along social connections, network-based sampling approaches, such as RDS, offer a promising approach for capturing these dynamics within communities of PWSD (6). This may be especially relevant in rural communities, where social and geographic isolation can further influence network connectivity (14). Beyond substance use, other characteristics including racial population group, education, and housing status are among several factors that have also led to health inequity in South Africa (15). These divides, rooted in South Africa’s history of post-Apartheid structural inequity, reinforce ongoing social segregation, leaving networks of PWSD both socially and geographically isolated from health systems and research efforts (16).

RDS relies on strong assumptions, including network connectivity, recruitment patterns, accurate reporting of social network size, and the ability of samples to reach equilibrium across recruitment waves. Yet, these assumptions may not be met in practice in populations characterized by social marginalization and geographic clustering (4,6,8). Homophily, the tendency for people to associate with others who share similar characteristics, represents one indicator of potential sampling bias; for example, a cluster with strong within-group recruitment may become overrepresented in the sample (17). Social networks of PWUD exhibit substantial clustering by demographic, behavioral, geographic, and social characteristics, which may influence recruitment patterns in such peer-referral studies (18,19). Other factors like recruitment chain stability and differential recruitment across participant characteristics may also influence RDS validity. Evaluating these methodological features is essential for understanding the strengths and limitations of RDS for PWSD.

This study quantifies RDS recruitment dynamics for PWSD in the rural town of Worcester in the Western Cape Province, South Africa (20,21). We (1) identify participant characteristics associated with *recruitment success*, defined as the distribution of at least one of two coupons that resulted in peer enrollment, (2) examine patterns of recruitment homophily across participant attributes, and (3) visualize how seed characteristics influence the stabilization of participant attributes across recruitment waves. These findings aim to improve understanding of RDS recruitment among PWSD in a rural, high TB/HIV-burden area, and may inform future research and intervention design in similar settings.

## Methods

### Study population and eligibility

Participants were recruited from Worcester between April 2021 and October 2023 (20). Worcester is a large rural town (estimated population 125,000 in 2023) composed of several distinct neighborhoods (22). The site was selected for its high burden of TB and HIV along with high illicit drug use. A clinical trial conducted in the community reported a TB prevalence of 2.2% (95% CI 1.4-3.4) among healthy individuals without HIV or a recent history of TB (23). In the region, 9.0% (95% CI 7.3-11.2) of individuals are living with HIV (24). While population-level estimates of drug use are limited, methamphetamine and methaqualone are widely available in the area.

In addition to presenting a recruitment coupon from another participant, eligibility included age ≥ 15 years, local residence in one of the neighborhoods in Worcester, and self-reported methamphetamine or methaqualone use within a month of enrollment, confirmed by positive urine drug tests. Participants were ineligible if they were currently intoxicated or unable to provide written informed consent. More details on the study are described elsewhere (20).

### Participant recruitment

With a total enrollment goal of 750, recruitment began with 2 initial seeds recruited from the community with plans to add seeds as needed to reach the target sample size while maintaining proportional representation across neighborhoods. Seeds and subsequent recruits received two physical coupons to distribute to peers with whom they smoked methamphetamine and/or methaqualone. Eligibility was verified upon enrollment. Each coupon contained a unique code to link enrolled participants to their recruiter, enabling the tracking of recruitment chains and assessment of recruiter success. This referral process continued in successive waves until the sample size was met. More details of recruitment design, compensation, and seed selection are described elsewhere (20).

### Data collection and study variables

The original purpose of this study was to estimate the prevalence of TB disease among people who smoke methamphetamine and/or methaqualone (20). Sputum was collected for TB diagnostic purposes, and blood was collected for rapid HIV testing. Demographic information was collected through surveys, and medical history data were self-reported and confirmed with medical records. Behavioral variables, including substance use and psychosocial characteristics, were collected using validated self-report questionnaires, including the Alcohol, Smoking, and Other Substance Involvement Screening Tool (ASSIST) for drug use assessment and mental health, the Center for Epidemiologic Studies Depression Scale (CES-D) to screen for depression symptoms, and the Household Hunger Scale (HHS) for food insecurity. These instruments have demonstrated validity in South African populations, including studies involving marginalized populations (25–28). Standard scoring and cutoffs were applied. Self-reported network size was collected for RDS adjustments and as a measure of social connectivity. Additional tools for the evaluation of socioeconomic status and mental health were administered but were not the focus of this analysis (20).

### Analysis Procedures and statistical software

Recruitment success, defined as the distribution of at least one of the two study coupons that resulted in peer enrollment, was the primary outcome of this analysis. A binary variable classified participants as having no peer recruits (unsuccessful) or at least one peer recruit (successful). Descriptive statistics were stratified by recruitment success to assess differences in demographic and psychosocial characteristics. Bivariate associations were tested using semi-parametric randomization tests for continuous and binary variables, which account for the correlation inherent in RDS data (29,30). Given the exploratory nature of this analysis, p-values less than 0.10 were used to identify potential associations, to generate hypotheses for future study rather than drawing definitive inferences.

Preferential recruitment, or recruitment homophily, was calculated to assess the tendency for participants to recruit peers with similar characteristics relative to the distribution expected under random recruitment (8,17). Homophily estimates were derived from observed recruiter-recruitee pairs within recruitment chains (e.g. the tendency for males to recruit other males). A homophily ratio close to 1 indicates little evidence of preferential recruitment for a given variable, whereas ratios greater than 1 indicate recruitment among similar individuals. Participant characteristics with ratios above 1.3 were flagged as demonstrating high homophily, which is consistent with previous work (31).

For recruitment chains that contributed more than 20 participants, sample proportions for key demographics across recruitment waves were analyzed visually to assess chain’s ability to diversify toward equilibrium as recruitment continued (17).

All analyses were conducted in R version 4.4.1using packages for RDS-specific estimation and association testing (32).

### Institutional Review Board Approval

The South African Medical Research Council (Pretoria, South Africa), Boston Medical Center (Boston, MA, USA), and Stellenbosch University (Tygerberg, South Africa) provided ethical approval for the study. All participants provided written informed consent.

## Results

### Recruitment overview

The parent study recruited 750 eligible PWSD; 749 were analyzed due to one participant’s missing self-reported network size. Ten seeds with confirmed methamphetamine and/or methaqualone use – 7 (70%) male, median age of 32 (IQR 27.0-35.5), 3 (30%) with current TB, and 2 (20%) living with HIV – were enrolled over the course of the study to reach the target sample size and initiate recruitment chains to improve representative of the community. Eight (80%) seeds successfully recruited peers to begin a chain, with 4 (40%) chains surpassing 20 participants. Among the 8 recruiting seeds, chains reached a median of 18.5 (IQR 8.8-52.5) participants across a median 7.5 (IQR 4.8-15.0) waves. The largest chain recruited 550 participants across 67 waves. Of the 749 participants, 499 (67%) recruited at least one peer; 258 (34%) who recruited one peer and 241 (32%) who recruited the maximum of two. Median self-reported network size recorded was 20 peers (IQR 10-50).

### Participant characteristics

Participant demographics, stratified by recruitment success, are shown in Table 1. Overall, participants were predominantly male (n=527; 70%) with a median age of 33 (IQR 28.0-39.0) years. Population group composition of the sample was mixed ancestry (n=608; 81%) and Black African (n=133; 18%). Compared with the broader Worcester population, the sample included a somewhat higher proportion of Black African participants. Afrikaans was the most common spoken language, regardless of racial population group (n=622; 83%). Overall, 133 (18%) participants were confirmed as living with HIV, 42 (32%) of whom were newly diagnosed.

**Table 1:** Participant Demographics by Recruitment Success with Homophily.

| **Characteristic** | **Overall**<br>**N = 749** | **No recruits**<br>**N = 250** | **One or two**<br>**recruits**<br>**N = 499** | **p-value** | **Recruitment**<br>**homophily** |
| --- | --- | --- | --- | --- | --- |
| Age (yrs) | 33 (28, 39) | 32 (28, 38) | 34 (29, 39) | 0.220 | 1.02 |
| Age group |  |  |  | 0.518 | 1.21 |
| <25 | 104 (14%) | 39 (16%) | 65 (13%) |  |  |
| 25-34 | 311 (42%) | 109 (44%) | 202 (40%) |  |  |
| 35-44 | 243 (32%) | 74 (30%) | 169 (34%) |  |  |
| 45+ | 91 (12%) | 28 (11%) | 63 (13%) |  |  |
| Born Male | 527 (70%) | 180 (72%) | 347 (70%) | 0.503 | 1.14 |
| Population group |  |  |  | 0.521 | 1.36 |
| Black African | 133 (18%) | 48 (19%) | 85 (17%) |  |  |
| Mixed Ancestry | 608 (81%) | 198 (79%) | 410 (82%) |  |  |
| Other | 8 (1.1%) | 4 (1.6%) | 4 (0.8%) |  |  |
| Preferred spoken language |  |  |  | 0.757 | 1.27 |
| Afrikaans | 622 (83%) | 202 (81%) | 420 (84%) |  |  |
| isiXhosa | 92 (12%) | 35 (14%) | 57 (11%) |  |  |
| English | 33 (4.4%) | 12 (4.8%) | 21 (4.2%) |  |  |
| Other | 2 (0.3%) | 1 (0.4%) | 1 (0.2%) |  |  |
| Self-reported network size | 20 (10, 50) | 20 (7, 50) | 20 (10, 50) | 0.265 | 0.82 |
| Neighborhood |  |  |  | 0.809 | 1.88 |
| A | 114 (15%) | 38 (15%) | 76 (15%) |  |  |
| B | 505 (67%) | 165 (66%) | 340 (68%) |  |  |
| C | 130 (17%) | 47 (19%) | 83 (17%) |  |  |
| Housing status |  |  |  | 0.771 | 1.22 |
| Formal residence | 488 (65%) | 161 (64%) | 327 (66%) |  |  |
| Informal residence | 252 (34%) | 85 (34%) | 167 (33%) |  |  |
| No current fixed residence | 9 (1.2%) | 4 (1.6%) | 5 (1.0%) |  |  |
| Education < 9th grade | 483 (65%) | 164 (66%) | 319 (64%) | 0.604 | 0.99 |
| Unemployed ** | 670 (89%) | 215 (86%) | 455 (91%) | 0.032 | 1.01 |
| Ever Incarcerated * | 478 (64%) | 172 (69%) | 306 (61%) | 0.053 | 1.06 |
| Within past 2 yrs | 189 (40%) | 66 (38%) | 123 (40%) |  | 1.16 |
| Depression risk (CES-D) * |  |  |  | 0.066 | 1.01 |
| High risk | 481 (64%) | 149 (60%) | 332 (67%) |  |  |
| Low risk | 268 (36%) | 101 (40%) | 167 (33%) |  |  |
| Household hunger score (HHS) |  |  |  | 0.267 | 1.14 |
| Moderate to severe | 306 (41%) | 95 (38%) | 211 (42%) |  |  |

| Characteristic | Overall<br>N = 749 | No recruits<br>N = 250 | One or two<br>recruits<br>N = 499 | p-value | Recruitment<br>homophily |
| --- | --- | --- | --- | --- | --- |
| Little to none | 443 (59%) | 155 (62%) | 288 (58%) |  |  |
| Living with HIV | 133 (18%) | 43 (17%) | 90 (18%) | 0.799 | 1.06 |
| New diagnosis upon<br>enrollment | 42 (32%) | 16 (37%) | 26 (29%) |  | 0.93 |
| Current TB disease | 72 (9.6%) | 24 (10%) | 48 (10%) | 0.917 | 1.00 |
| History of TB disease | 230 (31%) | 71 (28%) | 159 (32%) | 0.342 | 1.02 |
| Tobacco use | 710 (95%) | 237 (95%) | 473 (95%) | 0.976 | 1.01 |
| Alcohol use | 459 (61%) | 146 (58%) | 313 (63%) | 0.243 | 1.07 |
| Cannabis use * | 479 (64%) | 149 (60%) | 330 (66%) | 0.086 | 1.06 |
| Methamphetamine use † | 693 (93%) | 232 (93%) | 461 (92%) | 0.853 | 1.07 |
| Methaqualone use † | 691 (92%) | 228 (91%) | 463 (93%) | 0.457 | 1.01 |
| Multiple substances<br>reported | 687 (92%) | 228 (91%) | 459 (92%) | 0.730 | 1.01 |
| Tobacco risk ‡ |  |  |  | 0.165 | 0.99 |
| Abstains/Low | 41 (5.5%) | 14 (5.6%) | 27 (5.4%) |  |  |
| Moderate/Severe | 708 (95%) | 236 (94%) | 472 (95%) |  |  |
| Alcohol risk ‡ |  |  |  | 0.200 | 1.11 |
| Abstains/Low | 523 (70%) | 185 (74%) | 338 (68%) |  |  |
| Moderate/Severe | 226 (30%) | 65 (26%) | 161 (32%) |  |  |
| Cannabis risk ‡ |  |  |  | 0.108 | 0.97 |
| Abstains/Low | 274 (37%) | 100 (40%) | 174 (35%) |  |  |
| Moderate/Severe | 475 (63%) | 150 (60%) | 325 (65%) |  |  |
| Methamphetamine risk ‡** |  |  |  | 0.019 | 1.06 |
| Abstains/Low | 75 (10%) | 29 (12%) | 46 (9.2%) |  |  |
| Moderate/Severe | 674 (90%) | 221 (88%) | 453 (91%) |  |  |
| Methaqualone risk ‡** |  |  |  | 0.014 | 1.02 |
| Abstains/Low | 121 (16%) | 47 (19%) | 74 (15%) |  |  |
| Moderate/Severe | 628 (84%) | 203 (81%) | 425 (85%) |  |  |

Values reported as n (%) for categorical and median (Q1, Q3) for continuous
† Current use of methamphetamine or methaqualone was an inclusion criterion
‡ Reported via ASSIST survey
\* Significant at $\alpha=0.10$ ; \*\* Significant at $\alpha=0.05$

Seventy-two (10%) had confirmed TB disease at enrollment with an additional 230 (31%) reporting a prior TB diagnosis.

Few differences were observed between successful and unsuccessful peer recruiters at the α=0.10 threshold used to identify potential trends (Table 1). Both groups reported a median self-reported network size of 20 peers (p=0.265). Successful peer recruitment was more common among participants who were unemployed (91% vs 86%; p=0.032) or screened at high risk for depression (67% vs 60%; p=0.066), while a history of incarceration was associated with lower recruitment (61% vs 69%; p=0.053). Current cannabis use was more common among recruiters than non-recruiters (66% vs 60%; p=0.086), as was moderate-to-severe methamphetamine (p=0.019) and methaqualone (p=0.014) use based on ASSIST scores for these substances. Additional bivariate comparisons are shown in Table 1.

### Recruitment homophily

Two characteristics exceeded 30% likelihood to recruit similar peers (i.e. recruitment homophily ratio greater than 1.3): neighborhood of residence (1.88) and racial population group (1.36). Other characteristics that did not exceed 30% but showed considerable preferential recruitment were preferred spoken language (1.27), formal housing status (1.22), and age group (1.21), highlighting the tendency for participants to recruit peers with similar geographic and cultural backgrounds. There is little evidence of preferential recruitment by other participant characteristics, including TB (1.00) and HIV (1.06) (Table 1).

### Characteristic distributions

To assess how recruitment chains evolved over time, we examined the distribution of key participant characteristics across recruitment waves (Table 2). By wave five, all chains visually stabilized close to the sample’s overall proportion of males (n=527; 70%), regardless of the sex of the seed. The proportion of those with a history of incarceration stabilized around the sample proportion (n=478; 64%) in chains with seeds both with and without history themselves. Similarly, the proportion of participants with current TB disease leveled around the sample proportion of 10% for every chain. All four seeds of the high-recruiting chains tested negative for HIV, but the proportion of people with HIV still stabilized close to the sample proportion (n=133; 18%).

**Table 2:** RDS Recruitment Characteristics for Productive Chains.

| Seed | 2 | 3 | 9 | 10 |
| --- | --- | --- | --- | --- |
| with N > 20 | Female | Male | Male | Male |
|  | Incarc N | Incarc N | Incarc N | Incarc Y |
|  | TB- | TB- | TB- | TB+ |
|  | HIV- | HIV- | HIV- | HIV- |
|  | Net=10 | Net=20 | Net=39 | Net=50 |
| Wave reached | 67 | 27 | 9 | 11 |
| Chain N | 550 | 102 | 21 | 36 |
| Male % | 66.7 | 90.2 | 81.0 | 58.3 |
| Incarceration history % | 66.4 | 48.0 | 66.7 | 69.4 |
| Positive TB % | 10.7 | 4.9 | 4.8 | 11.1 |
| Positive HIV % | 18.0 | 15.7 | 38.1 | 8.3 |
| Self-reported network size (median) | 80 | 85 | 15 | 7 |

## Discussion

Hard-to-reach populations are often excluded from research due to lack of dedicated efforts toward their inclusion. Recognizing trends in their recruitment patterns can improve study design and efficiency, supporting both their inclusion in research and a more complete understanding of disease burden. With this analysis of a RDS study in Worcester, South Africa, we aimed to better understand how recruitment dynamics can inform both methodological considerations and intervention strategies for reaching underserved groups. While emerging research has begun to examine how participant characteristics influence recruitment patterns in RDS studies, little is known about PWSD and TB research, where high prevalence of drug use and continuing social inequities make recruitment patterns especially important to study (33–36).

We examined recruitment dynamics by assessing recruiter characteristics, recruitment homophily, and stabilization of participant traits across recruitment waves. Successful recruitment was more likely among participants who were unemployed, at risk of depression, or who reported cannabis use in the past three months. These associations may reflect greater availability of time, social network connectivity, and responsiveness to monetary incentives. Interestingly, although depression risk has previously been linked with lower research participation overall, our findings suggest that, once enrolled, participants at higher risk for depression were more likely to recruit peers (37). This association may reflect broader underlying psychosocial conditions correlated with depression risk, such as unemployment, substance use, or network-level characteristics that facilitate peer recruitment. History of incarceration was negatively associated with recruitment success, which may suggest disrupted social relationships or heightened distrust. The ASSIST scores for both methamphetamine and methaqualone use revealed positive associations between severity of use and recruitment success. Though use of either substance was an inclusion criterion for this study, this finding suggests that heavier use may lead to larger and more connected drug use networks, enabling recruitment ease. Previous United States-based RDS studies involving men who have sex with men found increased recruitment success in those with greater network size, but we found no such association among PWSD in this rural community (33,38). The predominance of male participants in this sample, consistent with the epidemiology of PWSD and TB, may indicate successful engagement of a population often underrepresented in comparable studies (39,40).

Using measures of recruitment homophily, we examined whether recruiters tended to recruit peers with similar characteristics. This preferential recruitment was observed across neighborhood of residence and race. Although collinearity was not formally evaluated, these characteristics are closely aligned within this setting and likely reflect overlapping social and spatial structures shaped by historical inequities (41). Similar patterns of homophily observed across these variables, including to a lesser extent primary spoken language, are not unexpected. These patterns suggest that recruitment occurs within tightly connected social groups, reflecting underlying network clustering. While such clustering may limit cross-group recruitment and challenge the extent to which the sample approximates a well-mixed population, these same network connections are what RDS leverages to facilitate peer-driven recruitment among hard-to-reach individuals. Interestingly, little evidence of recruitment homophily was observed for current TB disease, suggesting that TB was not strongly clustered within the observed recruitment network.

We assessed whether participant trait proportions stabilized across recruitment chains around the sample average regardless of seed characteristic. Consistent with RDS assumptions, we observed this stabilization in the four traits analyzed – sex, incarceration history, TB status, and HIV status – within 5 waves of recruitment (6,17). Notably, neither of the two chains initiated by seeds living with HIV reached 20 participants. While excluded from the stabilization analysis, this may reflect barriers to recruitment within this group, potentially related to additional stigma, differences in social network structure, or reduced willingness to recruit peers. Such challenges have been widely documented in HIV surveillance and peer-driven recruitment studies and may limit the ability of recruitment chains to propagate within this subgroup (42–45). Given that only two seeds in our study have HIV, these observations should be interpreted with caution.

While our results provide insights into RDS recruitment dynamics, some limitations should be considered. Given the exploratory nature of this analysis, no prior hypotheses were set, and no corrections for multiple comparisons were applied. While the significance of the bivariate associations should be interpreted with caution, they identify potential recruitment differences that warrant future investigation. Second, some recruitment chains did not advance beyond five participants. This limited wave progression restricts our ability to understand the individual, social, or structural barriers that may have prevented further peer recruitment, reflecting broader limitations of RDS methods that should be mentioned. RDS-adjusted measures like recruitment homophily rely on accurately reported network size, which are difficult to verify through self-report. The assumption that recruitment occurs randomly within social networks is also challenging to meet or confirm. In our study, evidence of homophily suggests that recruitment is often clustered within attributes, potentially limiting the spread across diverse groups. Finally, as with all RDS studies, the absence of a true sampling frame for hard-to-reach populations makes it difficult to determine the extent to which our sample fully represents the underlying population.

Our findings support RDS as a feasible and useful approach for engaging PWSD, a hard-to-reach population, and individuals affected by communicable diseases like TB and HIV. Insights into participant characteristics associated with recruitment success may inform the design of more efficient peer-recruitment strategies, such as tailoring coupon distribution based on recruiter attributes linked to successful recruitment. Future work could refine the definition of recruitment success by distinguishing between participants who recruit one versus two peers, relative to the number of coupons available. Additionally, further analyses could distinguish coupons distributed from those returned, since participants may distribute coupons without achieving peer enrollment, highlighting differences between recruitment effort and realized enrollments. Beyond surveillance, RDS holds potential for intervention-oriented research, including community screening for TB or HIV. By successfully reaching hard-to-reach communities of PWSD in this setting, this study illustrates the utility of RDS for both methodological advancement and public health interventions.

**Fig 1.**
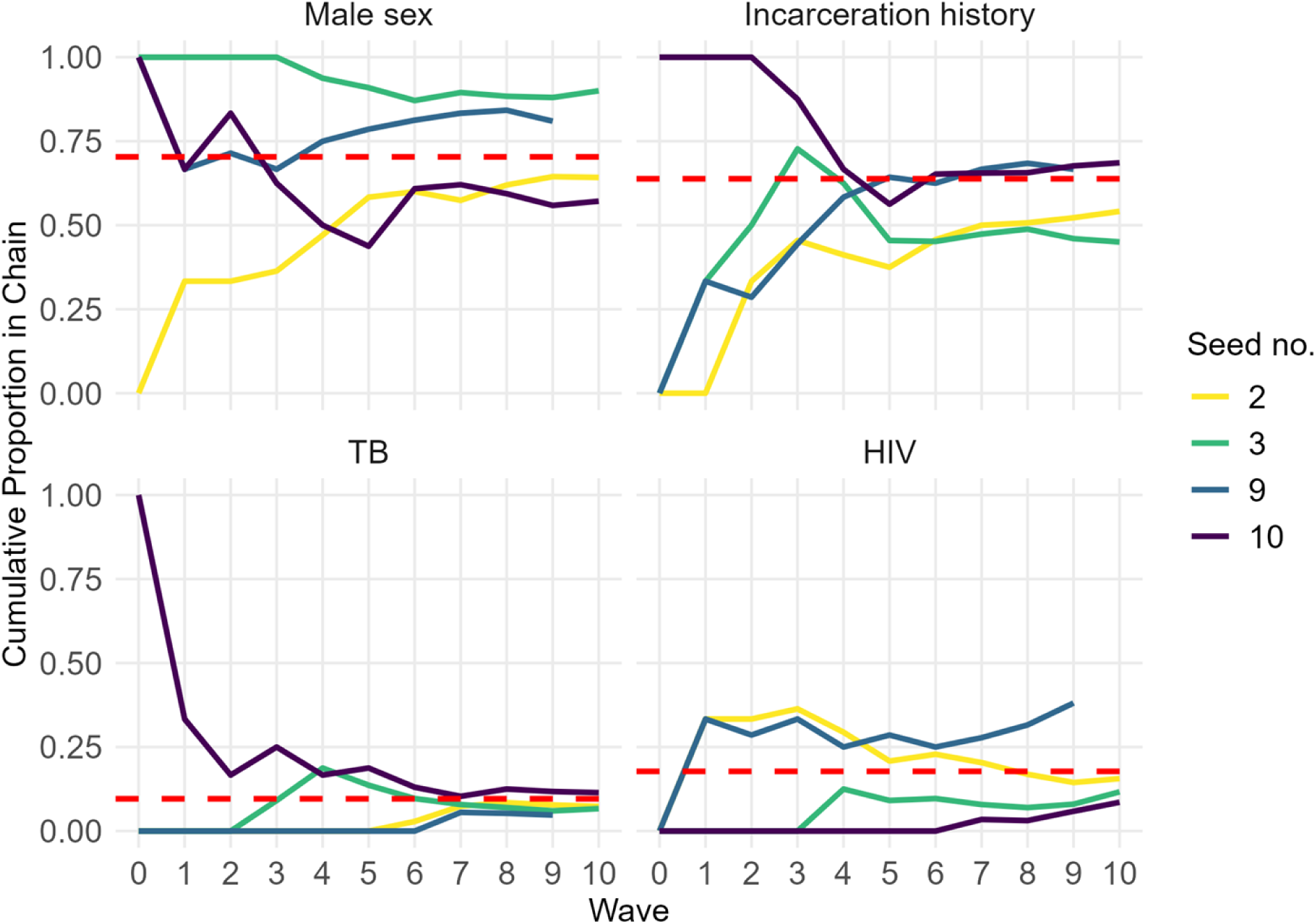
Stabilization of participant characteristics across recruitment waves for chains with n>20. Cumulative proportions of participants who are male, participants with a history of incarceration, participants with current TB disease, and participants living with HIV across recruitment waves. Each solid, colored line represents a recruitment chain initiated by a different seed. Only recruitment chains contributing more than 20 participants are shown. Dotted red lines represent the corresponding overall ample proportions (male = 0.70; incarceration history = 0.64; TB = 0.10; HIV = 0.28). The x-axis is limited to 10 recruitment waves for visualization, although some chains continued beyond wave 10.

## Data Availability

The data are available from the corresponding author upon request.

